# Advancing Research on Mechanisms of Resilience (ARMOR) Prospective Longitudinal Study of Adaptation in Young Military Recruits: Protocol and rationale for methods and measures

**DOI:** 10.1101/2023.07.07.23292348

**Authors:** Melissa A. Polusny, Craig A. Marquardt, Shelly Hubbling, Emily Hagel Campbell, Paul A. Arbisi, Nicholas D. Davenport, Kelvin O. Lim, Shumel Lissek, Jonathan D. Schaefer, Scott R. Sponheim, Ann S. Masten, Siamak Noorbaloochi

**Affiliations:** Minneapolis VA Health Care System, Minneapolis, MN; Center for Care Delivery Outcomes Research, Minneapolis, MN; Department of Psychiatry and Behavioral Sciences, University of Minnesota Medical School; Department of Psychology, University of Minnesota, Minneapolis, MN; Institute of Child Development, University of Minnesota, Minneapolis, MN; Department of Medicine, University of Minnesota Medical School

**Keywords:** study protocol, military personnel, longitudinal studies, resilience, adaptive behavior, stress, adversity, mechanisms, protective factors

## Abstract

**Background:** Military service provides a unique opportunity for studying resilience, a dynamic process of successful adaptation (i.e., doing well in terms of functioning and symptoms) in response to significant adversity. Despite tremendous interest in positive adaptation among military service members, little is known about the processes underlying their resilience. Understanding neurobiological, cognitive, and social mechanisms underlying adaptive functioning following military stressor exposure is essential to enhance the resilience of military service members.

**Objectives:** The primary objective of the Advancing Research on Mechanisms of Resilience (ARMOR) longitudinal study is to characterize trajectories of positive adaptation among young military recruits in response to Basic Combat Training (BCT), a well-defined, uniform, 10-week period of intense stress (Aim 1) and identify promotive and protective processes contributing to individual variations in resilience (Aim 2). The secondary objective is to investigate pathways by which neurobehavioral markers of self-regulation assessed by electroencephalography (EEG) and magnetic resonance imaging (MRI) contribute to adaptive trajectories (Aim 3).

**Methods:** ARMOR is an ongoing, prospective longitudinal cohort study of young military recruits who recently joined the National Guard but have not yet shipped for BCT. Participants (N=1,201) are assessed at five timepoints over the initial 2+ years of military service beginning before BCT (baseline) and followed up at 2 weeks, 6, 12, and 18 months post-BCT. At each time point, participants complete online questionnaires assessing vulnerability and protective factors, mental health and social-emotional functioning, and, at Time 0 only, a battery of neurocognitive tests. A subset of participants also complete structured diagnostic interviews, additional self-report measures, and perform neurobehavioral tasks before and after BCT during EEG sessions, and, at pre-BCT only, during MRI sessions.

**Results:** Study enrollment began April 14, 2019 and ended in October 16, 2021. A total of 1,201 participants are enrolled in the study (68.9% male; mean age = 18.9, SD = 3.0). Follow-up data-collection is ongoing and projected to continue through March 2024. We will disseminate findings through conferences, webinars, open access publications, and communications with participants and stakeholders.

**Conclusions:** Results are expected to elucidate how young military recruits adapt to military stressors during the initial years of military service. Understanding positive adaptation of military recruits in the face of BCT has implications for developing prevention and intervention strategies to enhance resilience of military trainees and potentially other young people facing significant life challenges.

## Introduction

Across the military career life cycle, service members are at considerable risk for exposure to stressors that may impact their health, well-being, and performance.^1–3^ Extensive research has identified factors that contribute to risk for psychopathology (e.g., post-traumatic stress symptoms, depression) following combat exposure and other military-related stressors. However, mounting evidence suggests that the majority of individuals show resilience, adapting successfully to risk and adversity.^4–8^ Understanding the neurobiological, cognitive, and social mechanisms underlying successful adaptation following military stressor exposure is essential to designing prevention and intervention strategies to enhance resilience for military populations.^9^ However, the mechanisms and processes that facilitate resilience following military stress remain poorly understood.

Most studies on resilience within the military context have operationalized resilience as a static, trait-like attribute or have relied on cross-sectional designs.^10^ One-time assessments of adaptation, adversity exposure, and resilience, particularly though self-report questionnaires, provide very limited insights into risk and protective processes and their influence on later mental health outcomes. There is now growing consensus that resilience is a multidimensional and dynamic process that unfolds over time in response to a challenge.^11–14^ We define resilience as the capability of a system to adapt successfully through multiple processes to challenges that threaten system function.^12^ Additionally, the resilience of an individual person draws on support from systems beyond the individual, including supportive relationships, such as battle buddies or unit support.^15,16^ Variations in the nature and timing of adversity exposure also influence how individuals adapt, and it is ideal to have assessments of adaptive functioning before, during, and following exposure to well-described uniform stressors. BCT provides a systematic and relatively uniform challenge with known timing, so that assessments of adversity, potential vulnerabilities, and protective influences, as well as adaptive functioning can be obtained before, during, and following a well-described period of challenge. Therefore, our approach to operationalizing resilience calls for a prospective, longitudinal study design that tracks the adaptation of military service members (MSMs) over time with assessments before and following BCT and related, well-defined challenges.

A growing body of longitudinal research with military populations has identified distinct latent classes (groupings) of service members demonstrating similar trajectories of adaptation over time.^5,17,18^ This literature suggests there is significant heterogeneity in people’s response to comparable stressors. However, these studies have generally examined trajectories following deployment and across the transition from military to civilian life.^19,20^ Little research has focused on young military recruits beginning at the outset of their military careers. During the initial years of military service, recruits are faced with numerous challenges (e.g., moving away from home, separation from family/friends, dramatic changes in living environment, and intense military training) as they are encultured into military life. Moreover, for most recruits, this transition from civilian to military life occurs within the context of also transitioning from adolescence to emerging adulthood.^21^ Identifying potential contributing factors linked to risk and resilience early in military service may be particularly useful in effectively intervening with service members during this key developmental transition. Longitudinal studies have also examined a limited number of outcomes in isolation (e.g., PTSD, depression, alcohol use). Our conceptualization of resilience as a multidimensional process, in which a person may show successful adaptation within some domains, but not others,^22^ calls for research that assesses multiple outcome domains (e.g., internalizing symptoms, externalizing problems, social-occupational functioning) over time. Finally, prior research has investigated the role of demographic characteristics (e.g., age, gender), psychological factors (e.g., neuroticism, self-efficacy), and environmental factors (e.g., social support, subsequent life stressors) in differentiating individuals who manifest resilience from those who exhibit maladaptive trajectories.^5,19,20^ Few studies have integrated neurobehavioral paradigms into longitudinal cohort studies,^17,23^ so the neurobiological, cognitive, and/or behavioral processes contributing to trajectory membership remains largely unknown.

Drawing on the above literature, the ARMOR study was established at the Minneapolis Veterans Affairs Health Care System (MVAHCS) and the University of Minnesota-Twin Cities (UMN).^24^ The overarching goal of this prospective, longitudinal cohort study with an embedded laboratory sub-study is to develop a comprehensive, multilevel model of resilience to guide the development of prevention and intervention strategies for military trainees and potentially other young people facing significant life challenges.

**Figure 1** presents the basic conceptual model guiding the study.^25^ It recognizes that individuals are embedded within a complex system of risk and protective factors that span multiple levels from individual neurobiological and behavioral factors to social and broader environmental factors. This multisystemic perspective acknowledges the dynamic and reciprocal relationships among these influences and their joint impact on resilience.^26^ The model also emphasizes potential pathways to risk and resilience in the face of adversity (shown as blue and red lines in response to BCT). Challenging life experiences past and present, including childhood adversity or trauma and exposure to military training, can heighten the risk of developing psychopathology. As illustrated by the dotted line in Figure 1, promotive factors, such as cognitive ability and social support, directly contribute to positive adaptation regardless of risk level. The dashed line in Figure 1 illustrates how protective factors can act as buffers against the detrimental effects of adversity. In the ARMOR study, the focus is on investigating self-regulation as a key protective process contributing to resilience.

**Fig. 1.**
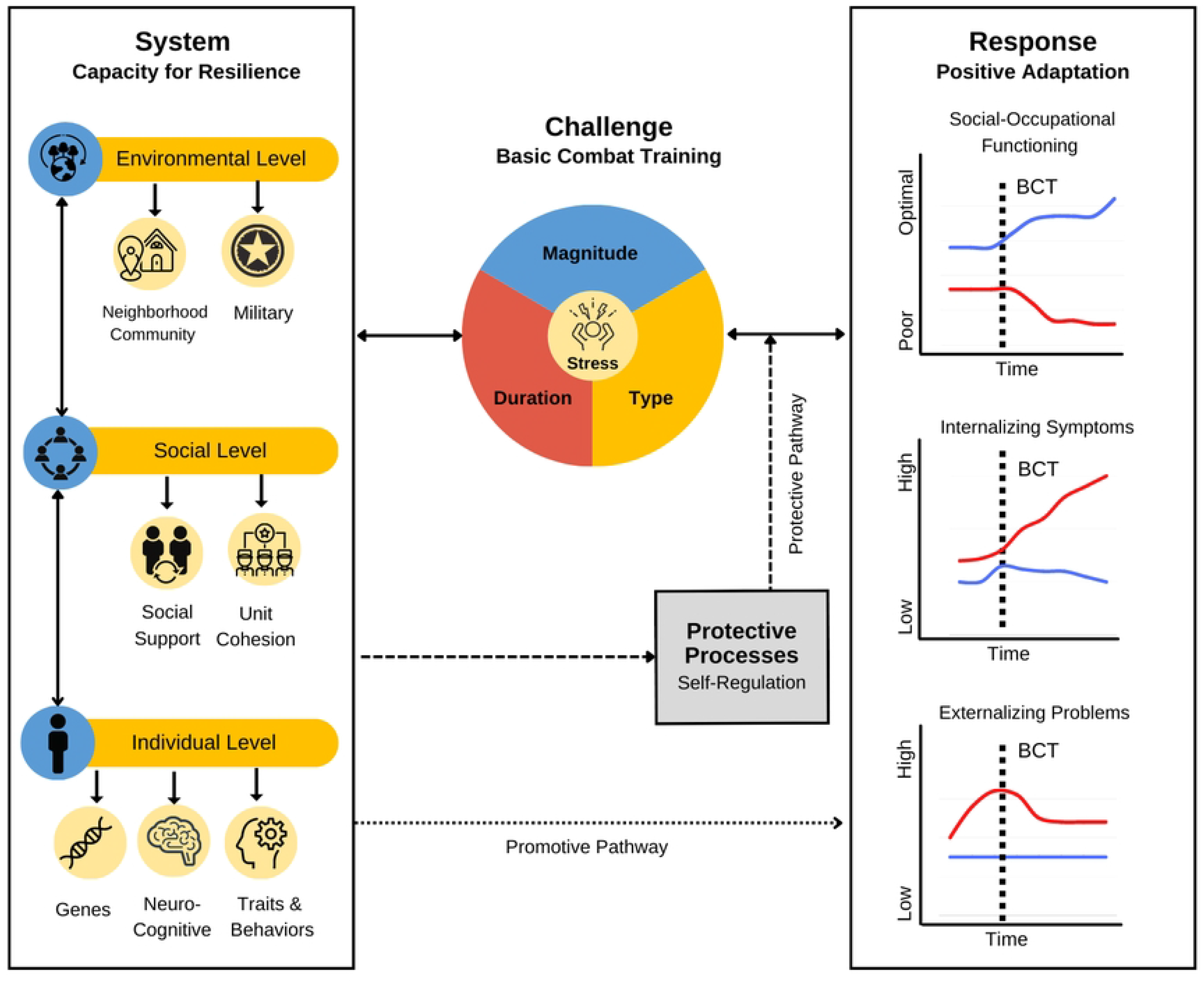
Integrated Multilevel Model of Resilience for Military Service Members *Note.* Conceptual model of resilience as a dynamic process in response to the challenges of Basic Combat Training (BCT). Risk and protective factors across multiple levels (individual, social, and environmental are depicted in the box on the left. Promotive and protective pathways are depicted with dashed and dotted lines, respectively. Positive (blue) and negative (red) responses to the challenges of BCT are shown in the graphs in the box on the right. From “A dynamic, multilevel approach to conceptualising and designing resilience research in the context of military stress,” MA, Polusny and CR, Erbes, 2023, *Stress & Health*, used under Creative Commons CC-BU license.

Self-regulation is comprised of three distinct but interrelated neurocognitive processes involving affect, behavior, and cognition, which together can facilitate adaptive responding to adversity.^27^ Attentional control involves the ability to concentrate and sustain attention despite distractions, inhibitory control refers to the regulation of maladaptive behavior in favor of goal directed actions, and behavioral flexibility enables the adaptation of behavioral strategies to meet environmental demands. Previous studies employing laboratory-based paradigms have identified significant associations between these self-regulatory processes and measures of trait resilience, implying their potential as stress-buffering mechanisms.^28^ However, few studies have examined neurobiological, cognitive, and/or behavioral processes in relation to resilience trajectory membership.^4^

### Study aims

The primary objective of the ARMOR study is to characterize trajectories of adaptation among young military recruits in response to BCT over the first two years of military service (Aim 1) and identify promotive factors and protective processes contributing to individual variations in adaptation (Aim 2). The secondary objective is to investigate whether neurobehavioral markers of self-regulation are predictive of resilient/non-resilient trajectories (Aim 3).

## Methods

### Study design

ARMOR is a prospective, longitudinal cohort study of young military recruits, including those who are age 17 at study entry, that investigates mechanisms contributing to manifested resilience across multiple levels of analysis (neural, cognitive, behavioral, and social). The study includes two components: (a) an online longitudinal survey component, including self-report measures presented via Qualtrics and neurocognitive tests using the Penn Computerized Neurocognitive Battery, and (b) a laboratory sub-study component, consisting of clinical diagnostic interviews, additional self-report questionnaires, and a series of performance-based tasks involving electroencephalography (EEG) and magnetic resonance imaging (MRI) assessments. We aimed to enroll a cohort of at least 1,200 young military recruits who recently enlisted in the Minnesota Army National Guard (MNARNG). Subject recruitment began on 4/14/2019 and ended on 10/16/2021. Data collection is ongoing and is expected to continue into 2024. As shown in **Figure 2**, participants are assessed at baseline (T0) before exposure to a uniform military challenge (BCT) and are currently being followed up at four time points (study is ongoing): two weeks following return from BCT (T1) and at six months (T2), 12 months (T3), and 18 months (T4) post-BCT. A subset of participants from the longitudinal cohort (n = 123) completed the laboratory sub-study procedures prior to shipping to BCT (pre-BCT) and after returning from training (post-BCT).

**Fig. 2.**
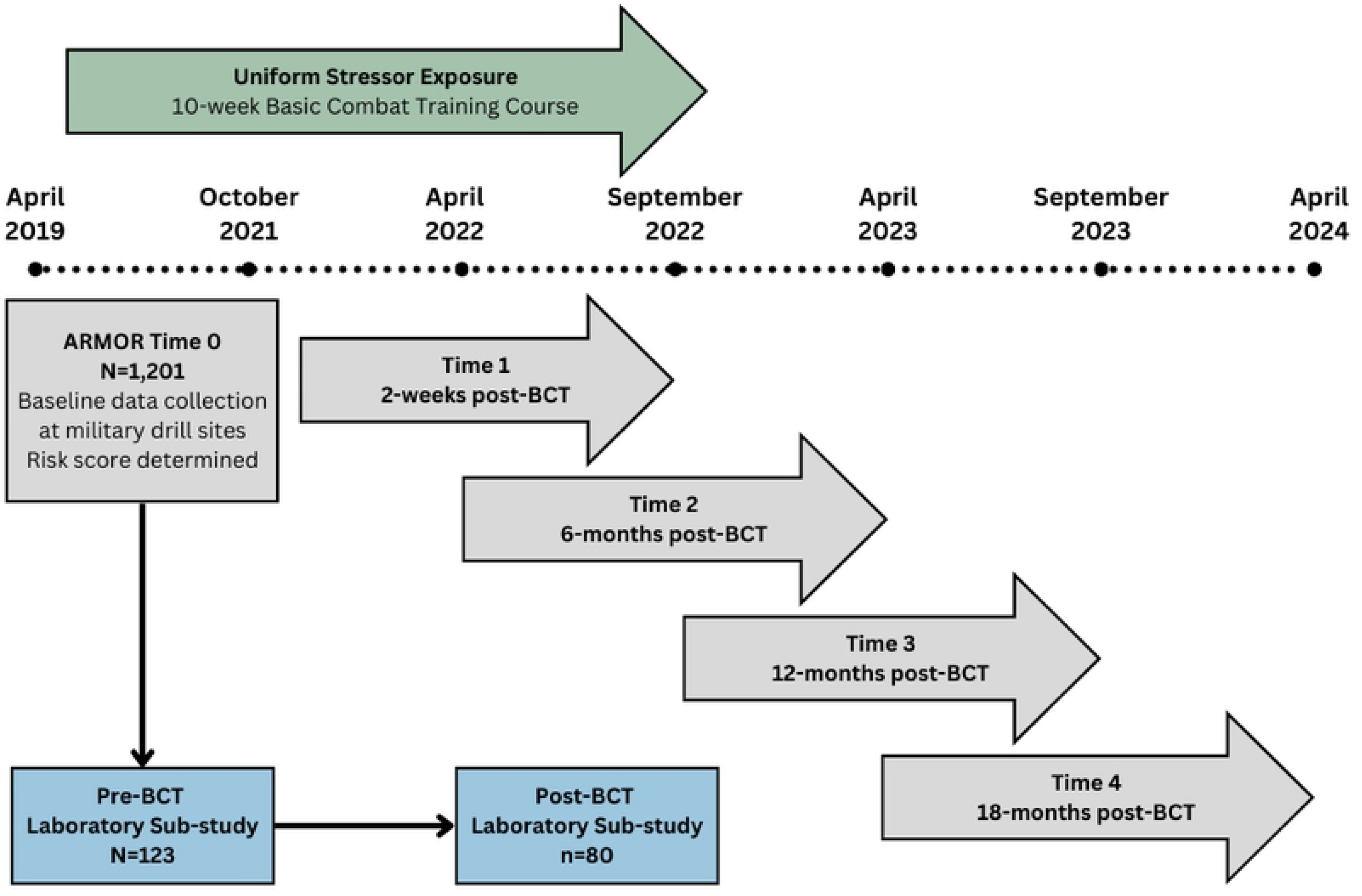

### Ethical considerations

This study has been approved by the Institutional Review Boards of the University of Minnesota (STUDY00004470) and Minneapolis VA Health Care System (VAM-18-00334/1594664). All study procedures were also approved by the relevant military command.

All participants provided informed consent. Participants were provided with a letter that explained all procedures, risks, and benefits. This information was verbally presented during briefings, and participants indicated their consent to take part in the survey component by entering a unique study ID number to begin the online survey. A waiver of the requirement to document consent for the survey component of this study was granted. The survey component of this study was also approved to include individuals under 18 years of age under 21 CFR 50.51/45 CFR 46.404. For the subset of participants 18 years or older taking part in the laboratory sub-study, written informed consent was obtained prior to participation in any laboratory study procedures.

### Context and setting

This study aims to investigate mental health and adaptive functioning among young military recruits before and after BCT, and beyond, offering a unique opportunity to study resilience in response to a well-defined, naturally occurring, uniform military stressor. BCT is a mandatory, intensive, 10-week training course that all military recruits must successfully complete. The training environment is highly structured, characterized by low personal autonomy, with strict discipline enforced by drill instructors.^29^ One distinctive aspect of BCT is the deliberate introduction of numerous stressors into the training environment, designed to prepare recruits for the challenges they will face in future military life.^30^ These challenges include prolonged separation/isolation from family/friends, dramatic changes in living environment, extreme physical demands, mental challenges, and simulated combat. This process is believed to foster unit cohesion and cultivate a strong sense of mental and physical toughness known as the “warrior ethos.” Previous research has shown that BCT has positive effects on cognitive performance, mood, and physical fitness,^31–35^ and it is associated with increased unit cohesion over time, which in turn, leads to reduced psychological distress and improvements in tolerating training stressors.^36^

While the majority of military recruits successfully adapt to the challenges of BCT, some individuals experience it as highly stressful and a considerable percentage (approximately 20%) of National Guard recruits fail to complete BCT.^37^ This suggests that although BCT is a uniform stressor experienced by all recruits, its impact and perceived stressfulness vary among individuals. A study of Swiss Armed Forces recruits participating in a 21-week BCT course found that higher perceived stress at the beginning of training was associated with greater mental distress and poorer military performance at later stages. Previous studies focusing solely on BCT completers have overlooked valuable data from a significant minority of individuals who exhibit reduced resilience.

To comprehensively understand how BCT influences adaptive functioning and study resilience processes over time, we employ a prospective, longitudinal research design with data collection before and after the relatively uniform stressor of BCT. This design makes it possible to capture change within subjects over time and allows us to test the temporal ordering of variables. Longitudinal designs permit stronger inferences about causal effects, although not the rigor of a randomized controlled trial. Our design also provides insight into the underlying mechanisms contributing to individual variations in resilience and allows for studying the impact of unforeseen events such as the COVID-19 pandemic and military deployments on resilience trajectories. By evaluating the perceived frequency and intensity of stressor exposure during BCT, we capture the individual experiences of military recruits. Furthermore, our focus on young, healthy participants at the beginning of their military careers, <u>before</u> significant exposure to military stressors, enables us to take a developmental approach to studying resilience within a military context. Finally, the integration of a laboratory sub-study within our research design provides an opportunity to conduct in-depth, multilevel assessments of a subset of the cohort. This laboratory sub-study allows us to investigate neurobehavioral markers of self-regulation that may predict resilience trajectories, enhancing our understanding of the underlying mechanisms associated with resilience in the context of military training.

### Participants and recruitment

Enrollment of participants for this study was based on specific inclusion criteria. Individuals aged 17 years or older, who were newly enlisted members of the National Guard and had been assigned a ship date to complete BCT during the study period, were eligible for participation. Exclusion criteria included a history of prior military service or any previous experience with BCT.

To recruit participants, a consecutive approach was employed within designated National Guard Recruitment Sustainment Program (RSP) units across the state until the target sample was reached. The research team conducted participant recruitment between April 14, 2019 and October 16, 2021, utilizing briefing presentations held during drill events at military posts. The relevant Army National Guard command provided the research team with a list of all potentially eligible MSMs. During briefings, interested individuals were provided with a study packet, which included a consent/assent letter, “Help Us Keep in Touch” locator form, “What to Expect Next” card illustrating the study design in layman’s terms, and a randomly generated unique study identification (ID) number. Throughout the recruitment process, detailed documentation was maintained. This included recording the total number of eligible MSMs approached, as well as the number of participants enrolled, those who refused to participate, and those found to be ineligible based on the inclusion and exclusion criteria.

### Survey data collection procedures

For the online survey component conducted at baseline (Time 0), participants were asked to complete a battery of self-report questionnaires on Qualtrics as well as select tests from the Penn Computerized Neurocognitive Battery (PCNB). Time 0 data was collected using study Chromebooks in classrooms at local armories. Participants were instructed to log into a secure online Qualtrics platform using their unique study ID for authentication. On average, the baseline data collection process took approximately 135 minutes (*SD* = 41).

Since exposure to BCT is required for continued follow-up, participants are monitored over the course of the study and retrospectively excluded if discharged prior to exposure to BCT. Participants’ exposure to BCT is verified prior to the initial follow-up wave (T1), and those eligible to continue study participation are invited to complete the battery of self-report questionnaires at each follow-up time point (see **Table 1**). All follow-up assessments are completed outside of drill training. Participants either complete a battery of computerized self-administered questionnaires using a confidential online survey which is linked to the participants’ study ID or via a paper-and-pencil version of questionnaires marked with study ID.

**Table 1.**
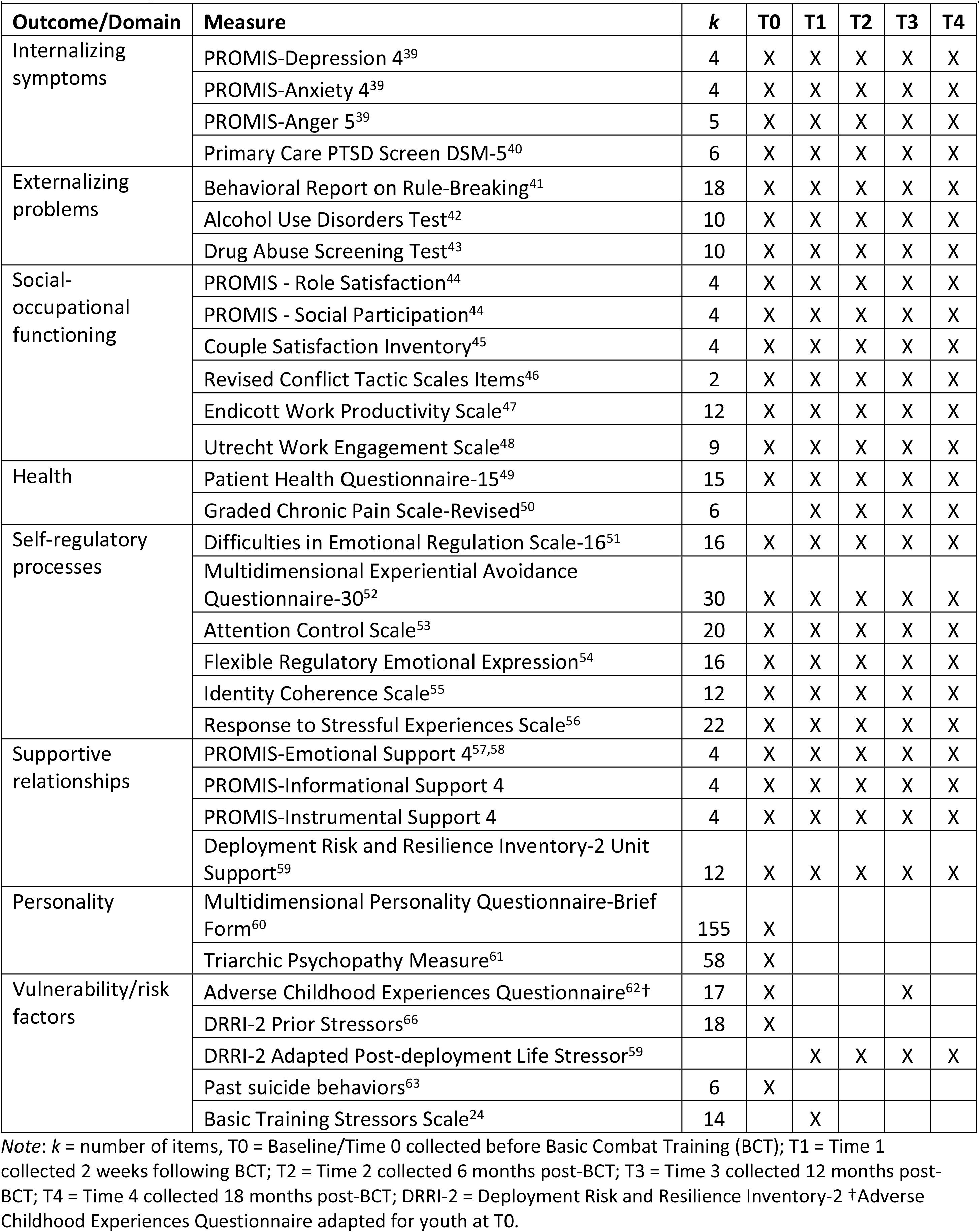
Self-report measures administered over time in the ARMOR longitudinal study

To ensure continuity of follow-up assessments, we adapted a longitudinal retention model developed by Scott and colleagues^38^ to engage and maintain contact with participants from enrollment through the final follow-up time point. During enrollment, participants were asked to provide detailed contact information, BCT ship and return dates, as well as full contact information for three alternate contacts who can help the study team locate the participant if needed. Study engagement will be maintained between follow-up waves through periodic communication (i.e., newsletters, greeting cards), and survey non-respondents will be contacted to encourage survey completion. Due to military regulations, participants did not receive financial compensation at baseline. However, potential participants were informed they would receive up to $150 for completing follow-up surveys.

### Measures

**Table 1** lists the battery of self-report questionnaires administered online via Qualtrics at baseline and at each follow-up. This battery of measures assesses eight domains (internalizing symptoms, externalizing problems, social-occupational functioning, health, self-regulation processes, supportive relationships, personality, and vulnerability/risk factors). While most questionnaires are repeated across all timepoints (T0-T4) as they are critical for tracking outcome trajectories and resilience processes, some are assessed only at T0 (i.e., personality) or T1 (i.e., BCT stressor exposure) as they do not require repeated assessment. **Table 2** provides a complete description of neurocognitive tests administered at baseline from the PCNB, which measures performance accuracy (e.g., proportion of correct responses) and speed (e.g., median correct response time) in major cognitive domains.^64^ All participants enrolled in the longitudinal study were also asked for a DNA saliva sample at baseline that is stored for future analysis.

**Table 2.** Tests from the Penn Computerized Neurocognitive Battery included in the ARMOR longitudinal study (Time 0 only)

| Neurobehavioral Function | Domain | Test |
| --- | --- | --- |
| Executive control | Attention | Penn Continuous Performance Test |
| Complex cognition | Language reasoning | Penn Verbal Reasoning Test |
| Episodic memory | Face memory | Penn Facial Memory Test |
| Social cognition | Emotion differentiation | Penn Emotion Differentiation Test |

**Table 3.** Self-report measures administered in the ARMOR laboratory sub-study pre-BCT and post-BCT

|  |  |  |  |
| --- | --- | --- | --- |
| **Measure** | ***k*** | **Pre-BCT Lab** | **Post-BCT Lab** |
| Patient Health Questionnaire-9<sup>73</sup> | 9 | X | X |
| Beck Depression Inventory-II<sup>70</sup> | 21 | X | X |
| Depressive Symptom Index — Suicidality Subscale<sup>71</sup> | 4 | X | X |
| State Trait Anxiety Inventory<sup>72</sup> | 40 | X | X |
| Positive and Negative Affect Schedule<sup>68</sup> | 20 |  |  |
| Anxiety Sensitivity Index<sup>73</sup> | 16 | X | X |
| Intolerance of Uncertainty Scale-Short Form<sup>74</sup> | 12 | X | X |
| *Note.* *k* = Number of items comprising measure, BCT = Basic Combat Training |  |  |  |

### Outcomes

Primary outcomes include trajectories of internalizing symptoms, externalizing problems, social-occupational functioning, and global adaptive functioning. Secondary outcomes are self-reported self-regulatory processes, neurobehavioral markers of self-regulation, and ecologically valid markers of functioning extracted from administrative military records.

### Administrative data

The study team will work with the local National Guard to extract administrative data capturing sociodemographic and service-related variables (i.e., Armed Forces Qualification Test score, training dates, and military discharge). Administrative data from military records will be de-identified and matched by participant ID.

### Laboratory sub-study data collection procedures

A subsample of the longitudinal cohort (n=123) was identified to participate in a laboratory visit both pre and post-BCT. Laboratory participants are selected based on their responses to baseline self-report measures and a predictive algorithm developed in the pilot UG3 phase of this project (details reported elsewhere).^65^ This strategy is intended to provide a subsample enriched with subjects at relatively high-risk of maladaptive functioning (n∼72) compared to a low-risk group (n∼48). Participants are briefly screened by phone, and those reporting contraindications to MRI due to the potential presence of metal in the body (e.g., employed as a metal worker, implanted medical devices) or due to problems with being in enclosed places are excluded.

Data for the laboratory sub-study is collected during an 8-9 hour combined total visit to the MVHCS and UMN Center for Magnetic Resonance Research (CMRR). First, participants undergo a structured clinical interview (1-2 hours) with a trained master’s level assessor under the supervision of a licensed clinical psychologist (CRE or PAA). The Structured Clinical Interview for DSM-5 is used to diagnose lifetime and current mental disorders.^66^ The Clinician Administered PTSD Scale for DSM-5 is used to diagnose PTSD.^67^ Next, while undergoing preparation for the EEG session, participants are asked to complete a battery of paper-and-pencil questionnaires specifically targeting constructs related to self-regulation (see **Table** 3).

### Electroencephalography (EEG)

Participants will complete an EEG session to measure resting state neural function as well as brain responses during cognitive tasks. We will use electrodes embedded in an elastic cap to record from 128 scalp sites. The precise physical location of electrodes will be recorded in three-dimensional space with respect to auricular and nasion landmarks so EEG recordings can be integrated with the corresponding structural MRI data. To measure eye movements for the detection of bioelectrical artifacts vertical electro-oculograms (VEOG) will be recorded from above and below the right eye and horizontal electro-oculograms (HEOG) will be recorded from outer ocular canthi. Left and right forearm electromyographs (EMGs), and electrocardiograms (EKG) will be recorded to identify and reduce artifact and quantify aspects of muscle contraction associated with button presses. EEG signals will be digitized at a rate of 512 Hz with 0.5 Hz low frequency cut-off and 200 Hz high frequency cut-off filters. Each participant will complete assessments of visual acuity and handedness as well as information about medications, alcohol, caffeine, and sleep in the last 24 hours. Self-report ratings of emotional state will be assessed before and after EEG recording sessions using the Positive Affect Negative Affect Scale (PANAS).^68^

During EEG collection at the pre and post-BCT visits, participants perform a series of neurobehavioral tasks assessing attentional control (dot probe paradigm),^75,76^ emotional inhibitory control (go/no-go paradigm),^77,78^ and feedback processing (gambling decision paradigm).^79^ **Table 4** provides a description of neurobehavioral tasks administered and domains assessed.

**Table 4.** Neurobehavioral tasks administered in the ARMOR laboratory sub-study

|  |  |  |  |  |
| --- | --- | --- | --- | --- |
| **Neurobehavioral task** | **Task description** | **Domain assessed** | **Pre-BCT** | **Post-BCT** |
| Dot Probe Task<sup>75</sup> | Participants passively inspect rapidly presented face pairs with neutral, happy, or angry/threatening expressions. Faces are replaced with either a blank space or an asterisk (target probe) placed in the same position as one of the two faces. Participants are instructed to respond as quickly as possible to indicate on which side the target probe appeared. | Attentional bias | X | X |
| Emotional Go/No-Go Task<sup>77</sup> | Participants view a series of rapidly presented visual targets and nontargets. Participants are instructed to press a response button for each target presented ("Go trials") and to avoid pressing the button for each nontarget ("NoGo trials"). Errors (response to NoGo trials or failure to respond to Go trials) are accompanied by visual (red bar in center of screen) and auditory (unpleasant buzzer sound) feedback. The task is completed in three blocks with adaptive response timing windows to increase task difficulty based on performance. Participants are also instructed they can win a monetary reward based on the number of points earned, which is displayed on screen every 20 trials. During the second block, time windows are shortened to increase the task's difficulty beyond what is feasible for participants. Points awarded during the second block for correct responses are also significantly reduced. | Inhibitory control | X | X |
| Gambling task<sup>79</sup> | For each trial, participants are presented with two adjacent squares each enclosing a number (5 or 25). Participants are instructed to choose between the two squares and then receive feedback (the chosen box turns red or green to signify either a win or a loss with red or green as the winning color counterbalanced across participants). | Error/performance monitoring, risk-taking | X | X |
| Farmer task<sup>80</sup> | The participant is a farmer whose task it is to travel between a shed and garden to harvest crops. Two different roads connect a shed to a garden: 1) a short road, and 2) a long road. Traveling the short road is perilous (contingently associated with electric shock [3-5mA, 100ms]) but allows the farmer quick travel from shed to garden and assures a successful harvest. Conversely, traveling the long road is always safe but often prevents the farmer from arriving at the garden before “wild birds” eat the crop. Through fear-conditioning, participants learn shock is predicted by a symbol, a ring of extreme size (CS+) but not the other size rings (conditioned safety-cue: CS-) displayed on the screen. The task measures fear generalization by assessing avoidance behavior in response to generalization stimuli similar but not identical to the CS+. | Maladaptive behavioral avoidance | X |  |

### Magnetic Resonance Imaging (MRI)

At the pre-BCT laboratory visit only, MRI data are collected at the University of Minnesota’s Center for Magnetic Resonance Research (CMRR) on a 3T Siemens Prisma scanner, using a 32-channel birdcage head coil with foam pads to minimize head movements. Sequences included T1-weighted MP-RAGE, T2-weighted SPACE, diffusion-weighted, resting state functional, and task-based functional MRI. While in the MRI environment, participants complete the Farmer task (see **Table 4**), a gamified task designed to assess adaptive versus maladaptive, avoidance responding in the face of physical threat. The task measures fear reactivity and behavioral avoidance to safe cues that resemble cues of imminent electric shocks.^80^ Decisions to ‘unnecessarily’ avoid during safe cues is considered *maladaptive* because danger (threat of shock) is not a realistic possibility, and avoiding unnecessarily compromises performance on the task. Adaptive responding is operationalized by higher rates of decisions to push through (i.e., low levels of avoidance) when encountering safe, yet danger-resembling, cues.

Finally, participants are asked to provide a blood sample to be stored for future research (30 min). Participants were compensated up to $400 for completion of both the pre-BCT and post-BCT visits.

### Data management

Data collected via the online survey platform (Qualtrics) administered at the UMN and will be downloaded securely into the study databases stored on a shared server at the MVAHCS with access limited to authorized study personnel. Qualtrics includes a complete suite of features to support HIPAA compliance, including a full audit trail, user-based privileges, and integration with the institutional LDAP server. Hard copy surveys will be double entered and checked for consistency. Laboratory sub-study data (i.e., neuroimaging and EEG data) will be stored in secure databases held on servers at UMN and are only available to lead investigators of each sub-study component. Administrative data will be transferred directly from MNARNG to a secure server at MVAHCS via secure data transfer. At MVAHCS, the data will be accessible by a named CCDOR Data/Statistics Team member. This individual will de-identify the personal information, match records with study ID number, and transfer it to shared server for the study. Study ID numbers will be used for data transfer, communication, and analysis purposes to protect confidentiality. Participant contact information linked with study ID number will be stored on a SQL server database will be created on internal VA web servers. Only individuals with a need to access the data, as vetted by the principal investigator are granted access. Access to contact information is obtained through Windows authentication (i.e., PIV card and password to the network).

#### Sample size and power analysis

Sample size calculations were based on the primary objective of the study (Aims 1-2) using structural equation models of various hypothesized structures (nested and longitudinal). This demand resulted in a sample size that could provide enough power to test such hypotheses across varying degrees of freedom. Control of Root Mean Square Error of Approximation (RMSEA) was the criteria used for our sample size and power calculations. According to MacCallum and colleagues,^81^ RMSEA= 0.01, 0.05, and 0.08 indicate excellent, good, and mediocre fit. **Table 5** gives the needed sample sizes and the corresponding powers for various degrees of freedom, assuming a null RMSEA of 0.05 and an alpha=0.05. Assuming a retention rate of 65% to reach an effective sample size of 780, we needed to recruit 1200 participants. Note that for n=780, models with df=5 and greater, would have an expected power of at least 0.909.

**Table 5.** Sample size and power calculations

| Sample size | df=2 | df=5 |
| --- | --- | --- |
| 700 | 0.567 | 0.878 |
| 800 | 0.619 | 0.916 |
| 1000 | 0.707 | 0.961 |
| 1200 | 0.776 | 0.982 |

#### Statistical analyses planned

Aim 1 will characterize adaptation among young military recruits over the first two years of service. Consistent with a hierarchical concept of the structure of resilience, we will use latent growth mixture modeling (LGMM) to identify the latent trajectories of adaptation and its indicators. We will apply LGMM at the level of internalizing symptoms, externalizing problems, and social-occupational functioning, as well as an overall model that simultaneously embeds all three domains into a model of adaptive functioning. We have hypothesized that at least three trajectories (one showing resilient adaptive functioning or low pathology, and others showing new onset and chronic distress) will emerge for each domain assessed (internalizing symptoms, externalizing problems, social-occupational functioning, and global adaptive functioning). <u>Note</u>: All remaining analyses for Aims 2 and 3 will be conducted on the trajectory class membership data for each of these four domain outcomes to test for specificity of resilience mechanisms.

Aim 2 will identify promotive factors and protective processes contributing to individual variations in adaptation. We will apply structural equation modeling and Bayesian Network Analysis to detect and identify the latent constructs and the dependency structure between different domains. We will also examine the effect of protective and vulnerability factors on membership in different trajectory classes. We propose to investigate this in two complementary ways. First, in our SEM modeling we will augment the measurement submodels of the most parsimonious latent growth structural mixture models accepted in Aim 1 with the indicators of protective/vulnerability measurements (**Tables 1 and 2**) and evaluate the change in the relationships (coefficients) indicators have with the latent trajectories. We will treat potential predictors as moderators or mediators, depending on the hypothesized role in the model, and we will test their contribution to the model by change in model fit indices (AIC, BIC, RMSE, G^2^). In our second approach, trajectory class membership assignment in Aim 1 will be used as dependent variables (resilience vs non-resilient/distressed trajectories) and protective/vulnerability factors as predictors in random intercept multinomial logistic mixed models that will provide a quantitative picture of the unique contribution of these measures. We hypothesize risk factors listed in **Tables 1 and 2** will predict membership in non-resilient/distressed trajectory classes, while protective/promotive factors will predict membership in the resilient/non-distressed trajectory class. All analyses will be estimated within the propensity classes that adjust for any possible lack of imbalance and will be performed both on the complete cases and five imputed data sets.

Aim 3 investigates whether neurobehavioral markers of self-regulation assessed using a series of performance-based tasks involving EEG and fMRI assessments are predictive of resilient/non-resilient trajectories. EEG recordings will undergo an independent component analysis-based processing pipeline to isolate and remove noisy time segments, signal artifacts (e.g., eye movements), and bad electrodes. Recording epochs centered around the onset of task stimuli will be extracted for the analysis of event-related potentials (ERPs) and time-frequency energy. Given the study focus on conflict monitoring and bottom-up covert attention, we plan to examine brain responses at midline frontal (e.g., FCz) and lateralized posterior electrodes (e.g., PO7, PO8), respectively. We will quantify brain response metrics such as feedback-related negativity and theta-band oscillations, which we predict will be associated with trajectory class membership. fMRI during the Farmer Task will be analyzed using an event-related design to model BOLD fluctuations to stimuli onsets using Analysis of Functional Neural Images (AFNI) software.^82^ Functional activation maps will be computed by regressing each voxel’s fMRI response time-course onto an ideal response function for 5 stimulus types: the danger cue, three classes of safety cues with parametrically varying levels of perceptual similarity to the danger cue, and a control condition including a safety cue with no similarity to the danger cue.^83^ We will focus particularly on fMRI responses to these stimuli in brain areas associated with fear reactivity (anterior insula, dorsomedial prefrontal cortex) and fear inhibition (ventromedial prefrontal cortex, anterior hippocampus). Fear-related brain activations to safe cues resembling danger cues (relative to safe cues without danger-cue resemblance) will be used as predictors of maladaptive avoidance (costly/unnecessary avoidance to safe cues resembling danger cues). Finally, these brain and behavioral responses, as well as their moderation by protective (e.g., social support, military cohesion, self-regulation) and vulnerability factors (e.g., BCT stressors, anxiety traits) will be used as predictors of trajectory class membership.

### Missing data

Every effort will be made to maximize complete data and responding across waves in the longitudinal survey (Aims 1 and 2). However, the collected data at each wave will be subject to various random missingness. At the end of each wave, the mechanism of missingness for each measured variable will be assessed and an imputation model proper to that variable (predictor/response/ adjustor covariate) will be identified. Appropriate imputation methods based on variable characteristics (e.g., regression for continuous variables, propensity or discrimination models or categorical models) will be used. After selecting a relevant model, and more importantly, deciding on the predictors of missingness, five imputed data sets will be constructed for each wave. The relevant analyses will then be based on the combined inferences derived from these five imputed data sets.

## Results

Study enrollment began April 14, 2019 and ended in October 16, 2021. A total of 1,201 participants are enrolled in the study (68.9% male; mean age = 18.9, SD = 3.0). Follow-up data-collection is ongoing and is projected to continue through March 2024.

## Discussion

To our knowledge, ARMOR is the first study to characterize trajectories of adjustment (i.e., resilient versus non-resilient) among US Army National Guard recruits using multidimensional assessments undertaken beginning at career onset and spanning two subsequent years. By assessing recruits before BCT and following up across 4 timepoints, the design provides an opportunity to map individual trajectories in response to exposure to a naturally occurring, uniform military challenge. The study aims to characterize adaptation among young military recruits over the first two years of service and identify processes contributing to individual variations in adaptation. While studies have identified numerous resilience factors, this study takes a longitudinal, multi-level approach to examine mechanisms underlying individual variations in resilience trajectories. In addition, this study utilizes an embedded laboratory sub-study design that integrates novel laboratory-based experimental paradigms for assessing dynamic self-regulatory processes into a large, prospective, longitudinal study. This approach will enable us to explore whether neurobehavioral markers of self-regulation assessed using a series of performance-based tasks involving EEG and fMRI assessments are predictive of resilient/non-resilient trajectories. By understanding the mechanisms underlying self-regulatory processes implicated in resilience, findings from our study will provide a foundation for the development of prevention and intervention strategies which may help to promote positive adaptation and resilience among young military recruits. Specifically, findings of this study may support the optimization of mindfulness-based interventions, which focus on increasing awareness of one’s thoughts, emotions, and actions to improve specific aspects of executive functioning including attention, cognitive control, and emotion regulation.

### Patient and public involvement

Military stakeholders from the local National Guard command were involved in the research process beginning at the early planning stage by identifying relevant research questions and providing consultation on feasibility of study methods. Outcome measures were informed by their priorities and military experience. Local National Guard command also facilitated the study team’s access to military personnel and allocated training time for the investigators to present information about the study to potential participants and collect baseline data.

Findings of the study will be shared with stakeholders through regular briefings to military command. Findings will be communicated to the public through various types of media, including print (e.g., newsletters, white papers) and broadcast channels (e.g., television and radio). Findings will be disseminated to the scientific community through peer reviewed publications and presentations.

## Data Availability

All relevant data are within the manuscript and its Supporting Information files.

## Acknowledgements

The authors wish to acknowledge Dr. Christopher Erbes, who passed away on May 30, 2021. Dr. Erbes served as Multiple PI on this project and contributed to the overall design and conceptualization of the study.

The content is solely the responsibility of the authors and does not necessarily represent the official views of the Departments of Veterans Affairs, Army, or Defense or the National Institutes of Health (NIH).

We received funding from UH3AT009651 from the National Center for Complementary and Integrative Health (NCCIH) of the NIH. This work also was supported by resources and the use of facilities at the Minneapolis VA Health Care System, Minneapolis, MN. We would like to express our gratitude to the National Guard service members volunteering to participate in the study.

## Authors’ Contributions

Melissa Polusny was responsible for the overall conception and design of the study and led manuscript writing.

Craig Marquardt made substantial contributions to the acquisition of data and revising the manuscript critically.

Shelly Hubbling made substantial contributions to the acquisition of data for the study, reviewing the manuscript.

Emily Hagel Campbell made substantial contributions to analysis and interpretation of data, reviewing the manuscript.

Paul Arbisi made substantial contributions to the acquisition of data and revising the manuscript critically.

Nicholas Davenport made substantial contributions to the acquisition of data and revising the manuscript critically.

Kelvin Lim made substantial contributions to the acquisition of data and revising the manuscript critically.

Shumel Lissek made substantial contributions to the acquisition of data and revising the manuscript critically.

Jonathan Schaefer contributed substantially revising the manuscript critically.

Scott Sponheim made substantial contributions to the acquisition of data and revising the manuscript critically.

Ann Masten contributed to the conception and design of the study, revising the manuscript critically.

Siamak Noorbaloochi contributed to the conception and design of the study, analysis and interpretation of data, and revising the manuscript critically.

All authors have provided final approval of the manuscript for submission.

